# Reported symptoms differentiate diagnoses in children with exercise-induced respiratory problems: findings from the Swiss Paediatric Airway Cohort (SPAC)

**DOI:** 10.1101/2020.03.12.20033340

**Authors:** Eva SL Pedersen, Carmen CM de Jong, Cristina Ardura-Garcia, Maria Christina Mallet, Juerg Barben, Carmen Casaulta, Karin Hoyler, Anja Jochmann, Alexander Moeller, Dominik Mueller-Suter, Nicolas Regamey, Florian Singer, Myrofora Goutaki, Claudia E. Kuehni

**Affiliations:** Institute of Social and Preventive Medicine, University of Bern, Switzerland; Division of Paediatric Pulmonology, Children’s Hospital St. Gallen, Switzerland; Division of Paediatric Pulmonology, Children’s Hospital Chur, Chur, Switzerland; Paediatric Pulmonology Practice Horgen, Horgen, Switzerland; Department of Paediatrics, University Children’s Hospital, University of Basel, Switzerland; Division of Paediatric Pulmonology, University Children’s Hospital Zurich, Switzerland; Department of Paediatrics, Kantonsspital Aarau, Switzerland; Division of Paediatric Pulmonology, Children’s Hospital Lucerne, Switzerland; Paediatric Respiratory Medicine, Children’s University Hospital of Bern, University of Bern, Switzerland; PedNet, Children’s University Hospital of Bern, University of Bern, Switzerland

**Keywords:** Exercise-induced, ILO, asthma, EIB, childhood, adolescents, dysfunctional breathing

## Abstract

**Background:** Exercise-induced breathing problems with similar clinical presentations can have different aetiologies. This makes distinguishing common diagnoses such as asthma, extrathoracic and thoracic dysfunctional breathing (DB), insufficient fitness, and chronic cough difficult.

**Objective:** We studied which parent-reported, exercise-induced symptoms (EIS) can help distinguish diagnoses of EIS in children seen in respiratory outpatient clinics.

**Methods:** This study was nested in the Swiss Paediatric Airway Cohort (SPAC), an observational study of children aged 0-17 years referred to paediatric respiratory outpatient clinics in Switzerland. We studied children aged 6-17 years and compared information on EIS from parent-completed questionnaires between children with different diagnoses. We used multinomial regression to analyse whether parent-reported symptoms differed between diagnoses (asthma as base).

**Results:** Among 1109 children, EIS were reported for 732 (66%) (mean age 11 years, 318 of 732 [43%] female). Among the symptoms, dyspnoea best distinguished thoracic DB (relative risk ratio [RRR] 5.4, 95%CI 1.3-22) from asthma. Among exercise triggers, swimming best distinguished thoracic DB (RRR 2.4, 95%CI 1.3-6.2) and asthma plus DB (RRR 1.8, 95%CI 0.9-3.4) from asthma only. Late onset of EIS was less common for extrathoracic DB (RRR 0.1, 95%CI 0.03-0.5) and thoracic DB (RRR 0.4, 95%CI 0.1-1.2) compared with asthma. Localisation of dyspnoea (throat vs. chest) differed between extrathoracic DB (RRR 2.3, 95%CI 0.9-5.8) and asthma. Reported respiration phase (inspiration or expiration) did not help distinguish diagnoses.

**Conclusion:** Parent-reported symptoms help distinguish different diagnoses in children with EIS. This highlights the importance of physicians obtaining detailed patient histories.

**Highlights box:**

1. What is already known about this topic? Experts suggest that information about the symptoms and their onset and duration can assist accurate diagnosis of children with exercise-induced respiratory problems, but no original studies have tested this. (29/35 words)
2. What does this article add to our knowledge? Exercise-induced symptoms reported by parents and further information about their onset, triggers, and effects of treatment help differentiate diagnoses in children with exercise-induced respiratory problems. (25/35 words)
3. How does this study impact current management guidelines? Our results emphasize the importance of taking detailed symptom histories of children with exercise-induced problems, and suggest which questions are most helpful.

## Introduction

Exercise-induced respiratory symptoms (EIS) are common in childhood. But their underlying causes can be difficult to identify because the clinical presentation of EIS of different aetiologies can overlap (1-3). EIS are most often caused by exercise-induced bronchoconstriction, extrathoracic or thoracic dysfunctional breathing (DB), insufficient fitness, and nonspecific chronic cough (4, 5). Even though clinical presentations can be similar, certain symptoms are typically associated with specific diagnoses (6-8). Knowing which symptoms are particularly characteristic of different underlying causes of EIS may facilitate diagnosis. For example, expiratory wheeze, cough, and shortness of breath are typical for exercise-induced bronchoconstriction with symptoms lasting from minutes to hours that usually peak after exercise (9, 10). Inspiratory problems with stridor, throat tightness, and shortness of breath are more typical for extrathoracic DB with symptoms that last only a few minutes and peak during exercise (9, 11). Typical symptoms for thoracic DB are shortness of breath, sighing, dizziness, and symptoms can last from minutes to hours and peak during exercise (12). Tingling in fingers or lips is typical for thoracic DB with hyperventilation.

A few studies have examined the association of diagnoses with typical symptoms. However, EIS have been reported only for specific diagnostic groups such as children with asthma (13), or no more than two diagnostic groups have been compared (14-16). If we better knew which symptoms most usefully distinguish diagnoses, misdiagnoses in children with EIS such as extrathoracic DB misdiagnosed as asthma might be reduced (17-19). We therefore studied children visiting paediatric respiratory outpatient clinics in Switzerland to investigate which symptoms reported by parents are most useful to distinguish different diagnoses of EIS.

## Method

### Study design

We used cross-sectional data from the Swiss Paediatric Airway Cohort (SPAC), a multicentre study of children referred to paediatric respiratory outpatient clinics in Switzerland (20). The SPAC study includes children aged 0-17 years who were referred for respiratory problems such as wheeze, cough, dyspnoea, or symptoms related to sleep or exercise, and who spoke sufficient German to participate. At the time of the visit, the physicians explained the SPAC study to the families. Parents completed a questionnaire before or shortly after the visit that inquired about symptoms, medication, environment, and health behaviours. After the visit, the SPAC study team collected the outpatient clinic letters that had been sent back to the referring paediatrician with information on diagnosis, diagnostic investigations, and treatment. We entered questionnaire responses and information from outpatient clinic letters into a Research Electronic Data Capture (REDCap) database (21). Recruitment for SPAC started in July 2017 and is ongoing. At the time we extracted data for this analysis, eight paediatric respiratory outpatient clinics in Switzerland were participating. Among 2971 children invited, 1838 (62%) agreed to participate (December 1, 2019).

The SPAC study was approved by the Bern Cantonal Ethics Committee (Kantonale Ethikkomission Bern 2016-02176). Written informed consent was obtained from parents and patients older than 13 years. This article follows the STROBE reporting recommendations (22).

### Inclusion criteria

We included children aged 6-17 years with a completed questionnaire and an available outpatient clinic letter with information on diagnosis. We restricted the population to schoolchildren because nearly all children referred for EIS to respiratory outpatient clinics are older than 5 years. The question used to identify children with EIS was “Does your child sometimes experience breathing problems during exercise?”

### Parent reported exercise-induced symptoms (EIS)

The questionnaire inquired about symptoms that included exercise-induced wheeze, cough, dyspnoea, tingling sensations in fingertips/lips, and other symptoms that could be reported in a free-text field. It also asked about characteristics of symptoms that included trigger factors (running, bicycle riding, intensive sport games, swimming), localisation of dyspnoea (chest, throat, or both), respiration phase (inspiration, expiration), onset of EIS (during or after exercise), duration of symptoms, and whether a short-acting bronchodilator helped to relieve symptoms. **Supplementary table 1** reproduces English translations of the questions about EIS in the German language questionnaire. Parental questionnaires were not disclosed to outpatient physicians.

### Diagnosis

Diagnosis was taken from the outpatient clinic letter that the hospital pulmonologists sent back to the referring physician. Some children were seen more than once in the outpatient clinic, and we took the diagnosis from the outpatient clinic with the latest date. We distinguished six diagnoses of EIS: asthma, extrathoracic DB, thoracic DB, asthma plus DB, chronic cough, and other (including insufficient fitness level, EIS of unknown aetiology, allergic rhinoconjunctivitis, recurrent respiratory infections, and rare pulmonary diseases). Exact definitions of diagnoses are in **Supplementary table 2**. If a child had more than one diagnosis listed in the letter, we used the diagnosis listed first, except in children who had asthma and any type of DB. In these children we created a separate category (asthma plus DB) because we believed that symptoms might differ between children with asthma, DB, and both occurring together. Outpatient clinic physicians were blinded to the parent completed questionnaire when giving the final diagnosis.

### Other variables

Age, sex, height, and weight were taken from the outpatient clinic letter. We calculated body mass index (BMI) as weight/height^2^ (kg/m^2^) and calculated age-adjusted BMI z-scores based on Swiss reference values (23), defining overweight as BMI z-score > 1. We obtained information on symptoms not induced by exercise from the questionnaire including parental education, environmental factors, and physical activity.

### Statistical methods

We compared proportions of EIS by diagnosis categories: asthma, extrathoracic DB, thoracic DB, asthma plus DB, chronic cough, and others using chi-square and Fisher’s exact tests. We studied which symptoms were most useful to distinguish diagnoses using multinomial logistic regression. We defined diagnosis as outcome and asthma as the reference category, and studied each explanatory EIS variable in turn, adjusted for age and sex. For the multinomial regression, due to the sample size we grouped chronic cough with other diagnoses. Overall, we had little missing information in the questionnaire replies (<7%) apart from the question about the respiration phase when EIS are worst (inspiration or expiration) for which 14% were missing. Children with missing data were excluded. We used STATA version 14 for statistical analysis.

## Results

Of the 1109 children aged 6-17 whose parents completed the questionnaire and for whom we had information about diagnosis, 732 (66%) reported EIS in the questionnaire (**supplementary figure 1**). On average, children with EIS were 11 years old (SD 3.2), 318 (43%) were female (**table 1**). Children with reported EIS were older and more often female than children without reported EIS. Among these children with EIS, 549 (75%) were diagnosed with asthma, 38 (5%) with extrathoracic DB, 30 (4%) with thoracic DB, 43 (6%) with asthma plus DB, 21 (3%) with chronic cough, and 51 (7%) received other diagnoses.

**Table 1.** Comparison of characteristics, respiratory symptoms and diagnoses between included patients with and without exercise induced symptoms (EIS) (N=1109)

|  | Yes to EIS in<br>questionnaire | No to EIS in<br>questionnaire |
| --- | --- | --- |
|  | N=732<br>n(%) | N=377<br>n(%) |
| Characteristics |  |  |
| **Demographic and socioeconomic characteristics** |  |  |
| Age (years), mean (SD) | 11.0 (3.2) | 9.5 (3.1) |
| Sex (female) | 318 (43) | 132 (35) |
| BMI z-score, mean (SD) (n=1091) | 0.3 (1.2) | 0.2 (1.2) |
| Overweight (BMI z-score >1) (n=1094) | 177 (26) | 76 (23) |
| Sports apart from at school (n=1056) | 566 (78) | 257 (70) |
| Swiss nationality | 609 (83) | 309 (82) |
| Parental education |  |  |
| Mother, tertiary<sup>a</sup> (n=1071) | 259 (37) | 114 (31) |
| Father, tertiary<sup>a</sup> (n=1056) | 315 (45) | 142 (39) |
| Parental smoking |  |  |
| Mother, current smoking (n=1090) | 114 (16) | 49 (14) |
| Father, current smoking (n=1046) | 174 (25) | 80 (23) |
| **Respiratory symptoms in the past 12 months** |  |  |
| Cough apart from colds, yes often (n=1096) | 88 (12) | 55 (15) |
| Cough at night apart from colds (n=1079) | 329 (46) | 151 (41) |
| Wheeze (n=1086) | 452 (63) | 168 (46) |
| >3 attacks of wheeze (n=1086) | 216 (30) | 48 (13) |
| Rhinitis apart from colds (n=1100) | 479 (66) | 213 (57) |
| Eczema ever (n=1090) | 215 (30) | 102 (28) |
| **Diagnosis given at outpatient clinic** |  |  |
| Asthma | 549 (75) | 276 (73) |
| Extrathoracic dysfunctional breathing | 38 (5) | 1 (0) |
| Thoracic dysfunctional breathing | 30 (4) | 7 (2) |
| Asthma + any DB | 43 (6) | 1 (0) |
| Chronic cough | 21 (3) | 35 (9) |
| Other | 51 (7) | 57 (15) |

<sup>a</sup> Degree from university of applied sciences or university. Abbreviations: EIS, exercise induced symptoms

Symptoms differed between diagnostic groups (**figure 1, table 2, figure 2**). Results from our multinomial regression analysis (adjusted for age and sex) showed that wheeze was reported less often for children with other diagnoses (relative risk ratio [RRR] 0.2, 95% CI 0.1-0.4) than for children with asthma. Cough was less common in children with thoracic DB (RRR 0.3, 95% CI 0.2-0.7) and asthma plus DB (RRR 0.3, 95% CI 0.2-0.6) than in children with asthma alone. Dyspnoea was reported more often for children with thoracic DB (RRR 5.4, 95% CI 1.3-23.1) and asthma plus DB (RRR 4.9, 95% CI 1.5-16.2) than in children with asthma alone. A tingling feeling in fingertips or lips was more common in children with thoracic DB (RRR 3.0, 95% CI 1.2-7.3) than in children with asthma.

**Table 2.** Reported exercise-induced symptoms by diagnosis group in children who reported exercise-induced respiratory symptoms in the questionnaire (n=732)

|  | Asthma | DB extra-<br>thoracic | DB<br>thoracic | Asthma +<br>any DB | Cough | Other | P-value |
| --- | --- | --- | --- | --- | --- | --- | --- |
| Baseline questionnaire | N=549<br>N (%) | N=38<br>N (%) | N=30<br>N (%) | N=43<br>N (%) | N=21<br>N (%) | N=51<br>N (%) |  |
| Type of EIS (n=732)<sup>a</sup> |  |  |  |  |  |  |  |
| Wheeze | 329 (60) | 20 (53) | 15 (52) | 27 (63) | 9 (43) | 9 (18) | <0.001 |
| Cough | 390 (71) | 21 (55) | 13 (43) | 17 (40) | 18 (86) | 32 (63) | <0.001 |
| Dyspnoea | 376 (68) | 32 (84) | 28 (93) | 40 (93) | 10 (48) | 29 (57) | <0.001 |
| Tingling feelings in finger or<br>lips | 53 (11) | 6 (16) | 8 (30) | 10 (24) | 0 (0) | 10 (21) | 0.002 |
| Other symptoms | 50 (9) | 5 (13) | 7 (23) | 7 (16) | 4 (19) | 8 (16) | 0.016 |
| Trigger activities (n=697) |  |  |  |  |  |  |  |
| Run short (50-100 m) | 327 (63) | 27 (71) | 19 (66) | 29 (69) | 14 (70) | 34 (71) | 0.730 |
| Run far (>1 km) | 402 (77) | 32 (84) | 22 (76) | 35 (83) | 13 (65) | 35 (73) | 0.522 |
| Cycle | 254 (50) | 27 (75) | 19 (68) | 23 (59) | 9 (50) | 24 (53) | 0.034 |
| Intensive sport games<sup>#</sup> | 396 (75) | 27 (71) | 23 (77) | 37 (86) | 13 (65) | 34 (71) | 0.453 |
| Swim | 162 (31) | 13 (34) | 17 (59) | 20 (48) | 8 (40) | 24 (50) | 0.002 |
| Localisation of dyspnoea (n=496 of 515<br>with dyspnoea)<sup>b</sup> |  |  |  |  |  |  |  |
| Chest | 189 (52) | 15 (47) | 13 (46) | 19 (50) | 3 (30) | 19 (70) | 0.187 |
| Throat | 47 (13) | 9 (28) | 6 (21) | 6 (16) | 3 (30) | 4 (15) |  |
| Chest and Throat | 125 (35) | 8 (25) | 9 (32) | 13 (34) | 4 (40) | 4 (15) |  |
| Respiration phase<sup>c</sup> (n=628) |  |  |  |  |  |  |  |
| Inspiration | 214 (46) | 19 (51) | 12 (44) | 27 (64) | 9 (60) | 17 (44) | 0.271 |
| Expiration | 45 (10) | 0 | 2 (7) | 2 (5) | 0 | 5 (13) |  |
| Inspiration and Expiration | 209 (44) | 18 (49) | 13 (48) | 13 (31) | 6 (40) | 17 (44) |  |
| EIS start<sup>d</sup> (n=677) |  |  |  |  |  |  |  |
| During exercise | 344 (69) | 36 (95) | 24 (86) | 33 (81) | 9 (43) | 29 (60) | <0.001 |
| After ending exercise | 156 (31) | 2 (5) | 4 (14) | 8 (19) | 12 (57) | 19 (40) |  |
| Duration of EIS<sup>e</sup> (n=648) |  |  |  |  |  |  |  |
| 1-2 minutes | 189 (37) | 13 (34) | 5 (20) | 14 (34) | 8 (38) | 21 (45) | 0.503 |
| 5-10 minutes | 268 (53) | 22 (58) | 14 (56) | 23 (56) | 10 (48) | 22 (47) |  |
| Longer than 10 min | 48 (10) | 3 (8) | 6 (24) | 4 (10) | 3 (14) | 4 (9) |  |
| Used asthma-spray before or<br>during exercise?<sup>g</sup> (n=712) | 41 (77) | 15 (39) | 14 (47) | 37 (88) | 13 (62) | 21 (43) | <0.001 |
| Effect of asthma-spray<sup>h</sup> (n=494 of 511 who used<br>asthma spray) |  |  |  |  |  |  |  |
| EIS disappear | 172 (43) | 2 (14) | 2 (14) | 9 (25) | 1 (8) | 3 (17) | * |
| EIS are reduced | 204 (51) | 8 (57) | 9 (64) | 18 (50) | 8 (62) | 11 (61) |  |
| No effect | 23 (6) | 4 (29) | 3 (21) | 9 (25) | 4 (31) | 4 (22) |  |

This table displays n(%) with column percentages.
<sup>a</sup>Which symptoms does your child have during exercise?
<sup>b</sup>If reported dyspnoea: Where is the sensation of symptoms felt the strongest?
<sup>c</sup>When are the symptoms worst?
<sup>d</sup>When do the symptoms begin?
<sup>e</sup>After ending the exercise, how long do the symptoms usually stay?
<sup>f</sup>Does your child sometimes get a tingling sensation in fingertips or around the mouth during the EIS?
<sup>g</sup>Has your child ever used an asthma inhaler during EIS?
<sup>h</sup>How well does this asthma inhaler help?

**Figure 1:**
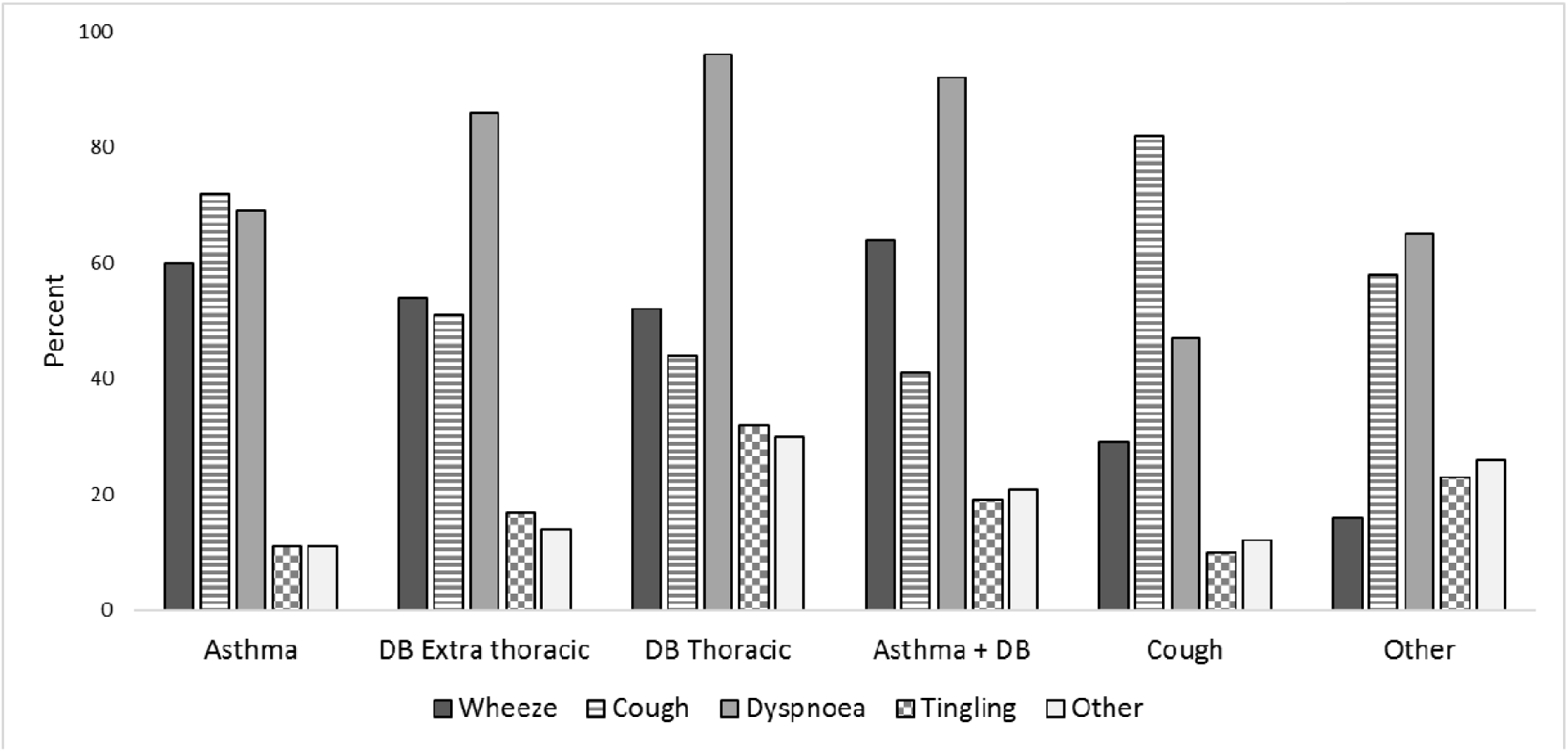
Type of exercise-induced symptoms in children referred to paediatric respiratory outpatient clinics, displayed by diagnosis group (n=732) DB, dysfunctional breathing

**Figure 2.**
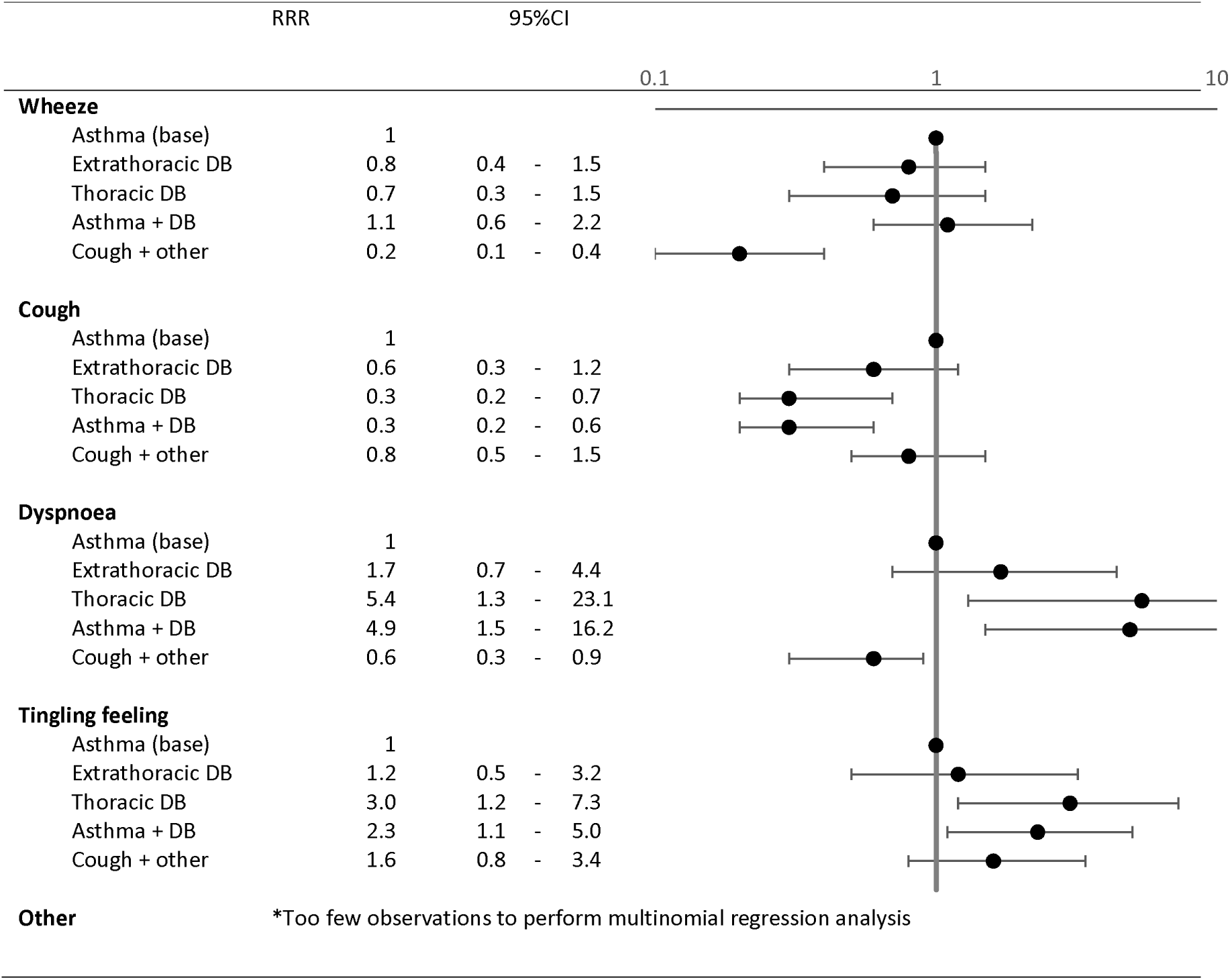
Comparison of type of EIS between diagnosis groups using multinomial regression models adjusted for age and sex. The graphs represent relative risk ratios from multinomial regression analysis with diagnosis categories as outcome (asthma as base variable) and types of symptom (wheeze, cough, dyspnea, tingling sensation in fingertips/lips, other symptoms) adjusted for age and sex. RRR, relative risk ratio; 95%CI, 95% confidence interval; DB, dysfunctional breathing

The type of physical activity reported to trigger EIS differed between diagnostic groups (**table 2, figure 3**). Compared to children with asthma, swimming was more commonly reported as trigger in children with thoracic DB (RRR 2.9, 95%CI 1.3-6.2), asthma plus DB (RRR 1.8, 95%CI 0.9-3.4), and other diagnoses (RRR 2.1, 95%CI 1.2-3.4). Bicycle riding was reported more often for children with extrathoracic DB (RRR 2.0, 95%CI 1.0-4.2), and intensive sports games were more often reported for children with asthma plus DB (RRR 2.4, 95%CI 1.0-5.8).

**Figure 3.**
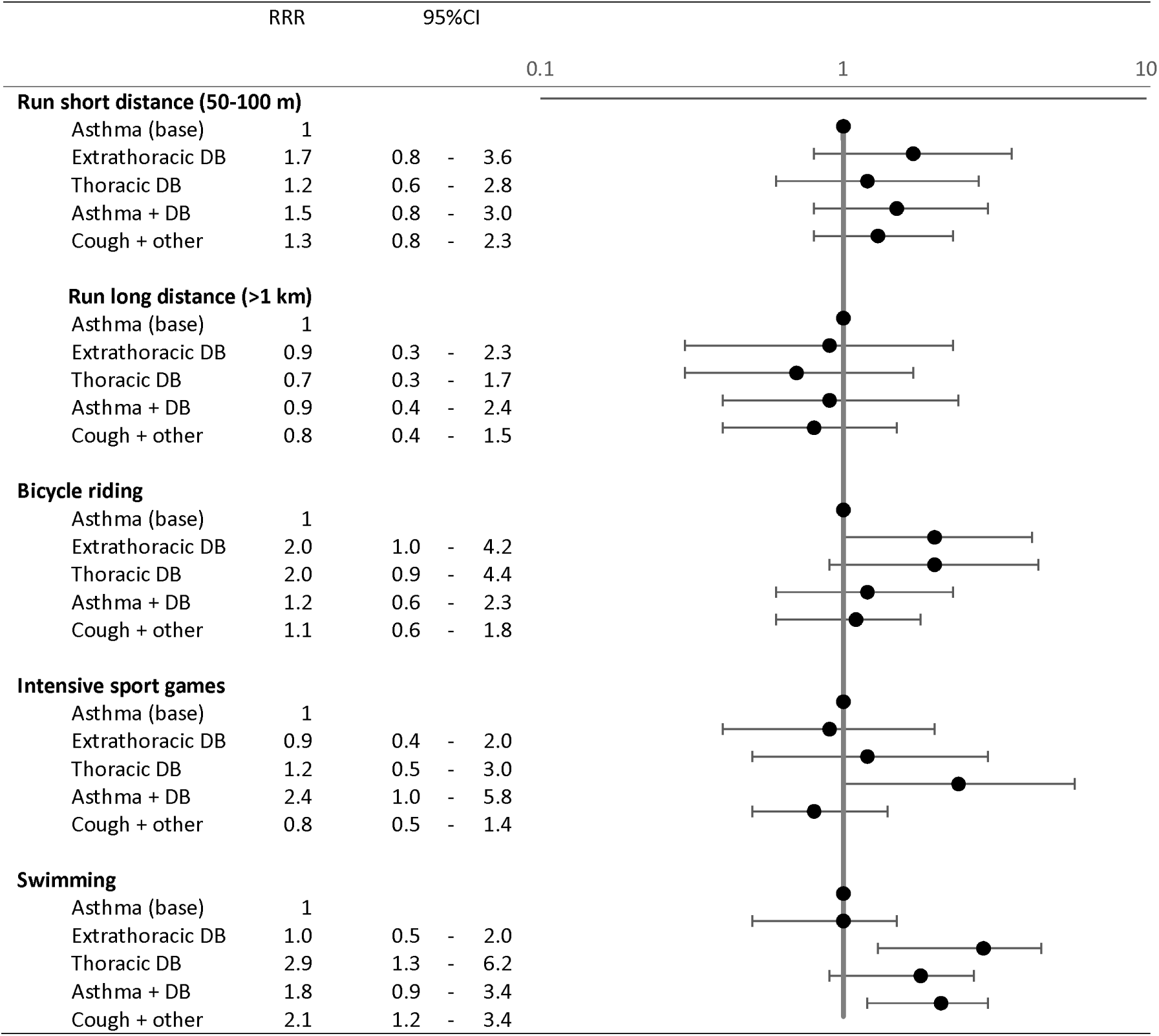
Comparison of activities triggering EIS between diagnosis groups using multinomial regression models adjusted for age and sex. The graphs represent relative risk ratios from multinomial regression analysis with diagnosis categories as outcome (asthma as base variable) and trigger activities (run, cycle, intensive sport games, swim) adjusted for age and sex. RRR, relative risk ratio; 95%CI, 95% confidence interval; m, meter; km, kilometre; DB, dysfunctional breathing

The characteristics of EIS differed between diagnostic groups (**table 2, figure 4**). Late onset (after exercise) of EIS was rarely reported for extrathoracic DB (RRR 0.1, 95% CI 0.03-0.5) compared to asthma. A long duration of EIS (more than 10 minutes) was reported more often for thoracic DB (RRR 4.8, 95% CI 1.4-16.8) than asthma. For localisation of dyspnoea, throat was reported more often than chest for children with extrathoracic DB (RRR 2.3, 95% CI 0.9-5.8) than for children with asthma. Respiration phase (inspiration or expiration) did not differ between diagnostic groups. Use of a bronchodilator made symptoms disappear in 172 (43%) children with asthma in contrast to 2 (14%) children with extrathoracic DB and 1 (8%) child with asthma plus DB (**table 2**).

**Figure 4.**
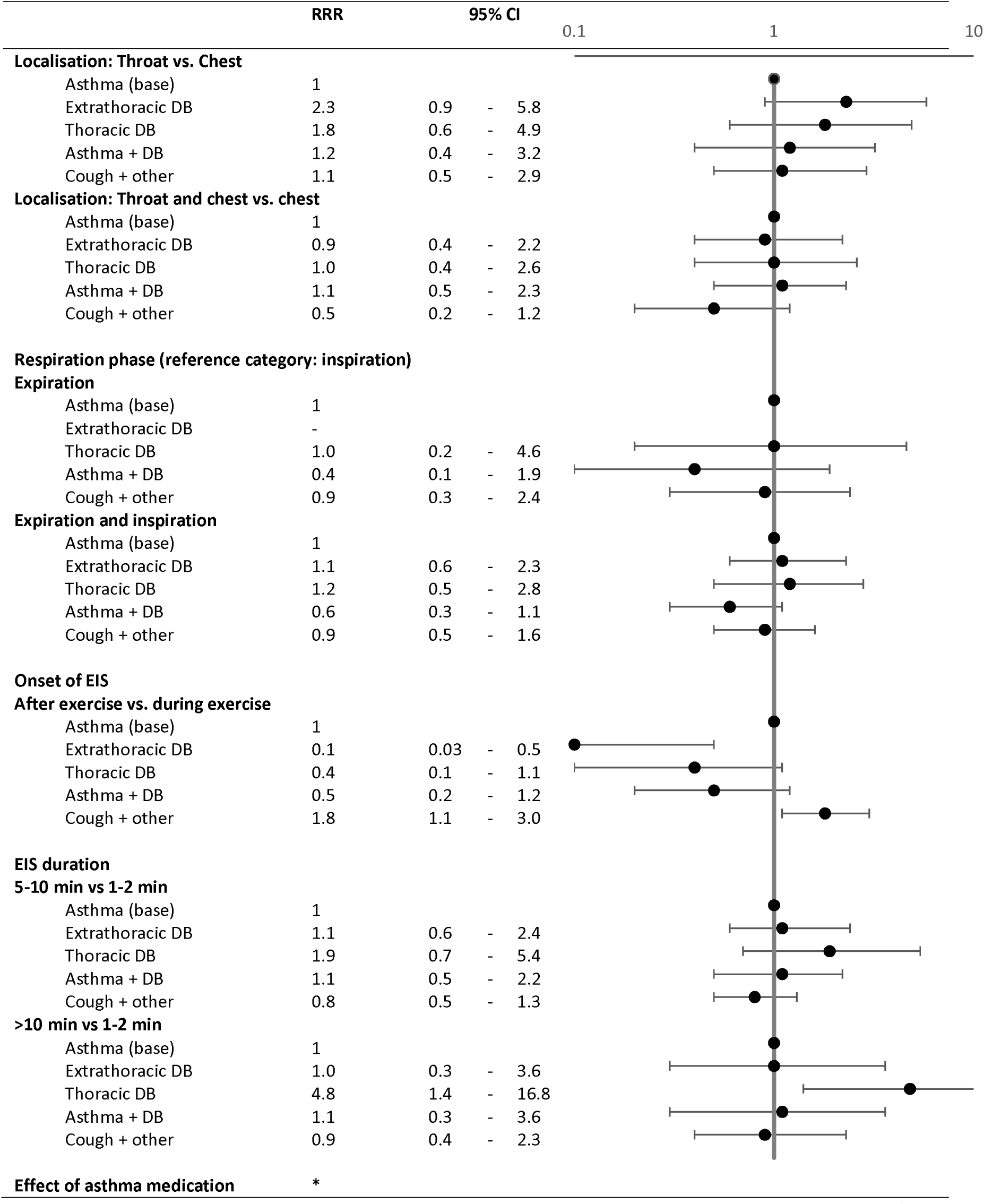
Comparison of EIS characteristics (localization of dyspnea, respiratory phase, onset, and duration of EIS) between diagnosis groups using multinomial regression models adjusted for age and sex. The graphs represent relative risk ratios from multinomial regression analysis with diagnosis categories as outcome (asthma as base variable) and characterisations of symptoms (localisation of dyspnea, respiration phase, duration of EIS) as explanatory variables. RRR, relative risk ratio; 95%CI, 95% confidence interval; DB, dysfunctional breathing *Too few observations to perform multinomial regression analysis

## Discussion

Parent-reported symptoms can distinguish different diagnoses in children with EIS referred to paediatric outpatient clinics. We observed that especially reported exercise-induced cough, dyspnoea, and tingling sensation in fingers or lips differed between diagnostic groups. Of the physical activities triggering EIS, intensive sport games and swimming best distinguished diagnosis groups. Additionally, onset of symptoms, duration of symptoms, and effect of a short-acting bronchodilator differed between the diagnostic groups. Respiration phase (inspiration or expiration) was less helpful.

### Strengths and limitations

Information about EIS and activities that trigger them has not previously been reported in such detail. The comparison of questionnaire-reported symptoms in children with EIS and diagnostic groups, which included asthma, extrathoracic DB, thoracic DB, and the combination of asthma and DB, is this study’s major strength. The level of detail afforded examination of how activities trigger different exercise-induced problems. In addition, our study was nested in SPAC, a real-life prospective observational clinical cohort which is representative of children referred to paediatric respiratory outpatient clinics for respiratory problems. We therefore believe our findings can be broadly generalised to children seen by respiratory physicians for EIS.

A limitation of the study is that we did not use results from diagnostic tests. Some tests (e.g., exercise challenge tests) were performed by indication and therefore not done in all children. Another limitation is that the questionnaire was addressed to the parents rather than the children themselves. However, we encouraged parents to fill in the questionnaire together with their child, which increases validity of reported symptoms (24, 25). Our questionnaire included the set of questions usually asked by physicians during the consultation. However, physicians might have worded some questions differently, addressed them to the child rather than the mother, simulated respiratory sounds such as wheeze or stridor, or provided additional explanations. So, although collected at the same time, the replies in the parental questionnaire, used for the analysis, might not always mirror the information retrieved by the physician who took the history. Our questionnaire did not include separate questions on expiratory wheeze and inspiratory stridor because there are no distinctive words in German for these two sounds. Most children with EIS, and also those with asthma, have the sensation that symptoms occur during inspiration rather than expiration (5). This might explain why we found no difference between diagnosis groups in whether symptoms were worse during inspiration or expiration; this question also had more missing answers. Our limited sample size for some diagnostic categories (thoracic DB, n=30) led to wide confidence intervals, and we could not investigate combinations of reported symptoms. Still, our study is the largest of its kind. A further limitation is that the final diagnosis was made by different pulmonologists and not based on a standardised, predefined diagnostic algorithm. However, all pulmonologists were board-certified and diagnoses were based on clinical history and standardized objective diagnostic test results representative of typical situations in a tertiary care clinic.

### Comparison with other studies

A few studies have presented questionnaire-reported symptoms for children or adolescents with EIS. A Swedish population based study in children aged 12-13 years reported exercise-induced symptoms for 128 children with an asthma diagnosis (13). Exercise-induced wheeze was reported for 76 (59%), cough for 81 (63%), and chest tightness for 56 (44%); throat tightness also was reported for 63 (49%), and inspiratory stridor for 47 (37%). We saw higher prevalence of symptoms overall because our study included respiratory outpatients and not children from the general population.

In a case series study of 12 adolescent athletes seen for suspected exercise-induced laryngeal obstruction (EILO) (15), dyspnoea during inspiration was reported by all (100%) and dyspnoea during expiration by 8 (67%), and throat tightness was reported more frequently (50%) than chest tightness (25%). A Danish study that compared 42 adolescents with EILO with 16 adolescents diagnosed with airway hyper-responsiveness (AHR) similarly found that all reported wheeze and stridor, but those with EILO mostly reported cough and throat tightness while those with AHR reported mostly dyspnoea (14). Our results and those from previous studies emphasize that no symptom is uniquely reported for single diagnostic groups among children with EIS, but some symptoms are reported more frequently for certain diagnoses than others.

### Interpretation

Cough, dyspnoea, and tingling sensation better distinguished thoracic DB from asthma than extrathoracic DB from asthma. This partly explains why extrathoracic DB can be misdiagnosed as asthma (17, 19). Onset of EIS during exercise was strongly associated with extrathoracic DB, while onset after exercise was associated with asthma. This finding is in line with the literature and could help physicians distinguish extrathoracic DB from asthma (7, 26, 27). We did not see any difference in the duration of symptoms between diagnostic groups, but parents may have had difficulty answering the duration question. Rewording the question in a validation study might help to find out if differences in duration exist.

Diagnosing children with EIS is not easy and requires a thorough diagnostic work up including objective diagnostic tests. Our study confirms that parent-reported symptoms can help to distinguish different diagnoses in children with EIS. This highlights the importance of physicians taking detailed symptom histories.

### Ethics approval and consent to participate

The SPAC study was approved by the Bern Cantonal Ethics Committee (Kantonale Ethikkomission Bern 2016-02176). Written informed consent was obtained from patients’ parents and directly from patients older than 13 years.

## Data Availability

The SPAC dataset is available on reasonable request by contacting Claudia Kuehni by.

## List of abbreviations

BMI: Body mass index
DB: Dysfunctional breathing
EIS: Exercise-induced symptoms
ILO: Inducible laryngeal obstruction
RRR: Relative risk ratio
SPAC: Swiss Paediatric Airway Cohort

## Author’s contributions

EP and CK made substantial contributions to the study conception and design. EP, CdJ, and MCM collected and prepared data from the SPAC study. EP drafted the manuscript. EP, CdJ, CA, MCM, JB, CC, KH, AJ, AM, DM, NR, FS, MG, and CK critically revised and approved the manuscript.

## Acknowledgements

We would like to thank the families who took part in the SPAC study. We would also like to thank the outpatient clinic assistants, nurses, and doctors for recruiting patients.

## Availability of data and material

The SPAC dataset is available on reasonable request by contacting Claudia Kuehni by.

## References

1. Connett GJ, Thomas M. Dysfunctional Breathing in Children and Adults With Asthma. Frontiers in pediatrics. 2018;6:406.

2. Johansson H, Norlander K, Berglund L, Janson C, Malinovschi A, Nordvall L, et al. Prevalence of exercise-induced bronchoconstriction and exercise-induced laryngeal obstruction in a general adolescent population. Thorax. 2015;70(1):57–63.

3. Fretzayas A, Moustaki M, Loukou I, Douros K. Differentiating vocal cord dysfunction from asthma. Journal of Asthma and Allergy. 2017;10:277–83.

4. Barker N, Everard ML. Getting to grips with ‘dysfunctional breathing’. Paediatric respiratory reviews. 2015;16(1):53–61.

5. Depiazzi J, Everard ML. Dysfunctional breathing and reaching one’s physiological limit as causes of exercise-induced dyspnoea. Breathe (Sheffield, England). 2016;12(2):120–9.

6. Kenn K, Hess MM. Vocal Cord Dysfunction: An Important Differential Diagnosis of Bronchial Asthma. Deutsches Ärzteblatt International. 2008;105(41):699–704.

7. Roksund OD, Heimdal JH, Clemm H, Vollsaeter M, Halvorsen T. Exercise inducible laryngeal obstruction: diagnostics and management. Paediatric respiratory reviews. 2017;21:86–94.

8. Roksund OD, Heimdal JH, Olofsson J, Maat RC, Halvorsen T. Larynx during exercise: the unexplored bottleneck of the airways. European archives of oto-rhino-laryngology : official journal of the European Federation of Oto-Rhino-Laryngological Societies (EUFOS) : affiliated with the German Society for Oto-Rhino-Laryngology - Head and Neck Surgery. 2015;272(9):2101–9.

9. Liyanagedara S, McLeod R, Elhassan HA. Exercise induced laryngeal obstruction: a review of diagnosis and management. European archives of oto-rhino-laryngology : official journal of the European Federation of Oto-Rhino-Laryngological Societies (EUFOS) : affiliated with the German Society for Oto-Rhino-Laryngology - Head and Neck Surgery. 2017;274(4):1781–9.

10. Chiang T, Marcinow AM, deSilva BW, Ence BN, Lindsey SE, Forrest LA. Exercise-induced paradoxical vocal fold motion disorder: diagnosis and management. The Laryngoscope. 2013;123(3):727–31.

11. Olin JT. Exercise-Induced Laryngeal Obstruction: When Pediatric Exertional Dyspnea Does not Respond to Bronchodilators. Frontiers in pediatrics. 2019;7:52.

12. Niggemann B. How to diagnose psychogenic and functional breathing disorders in children and adolescents. Pediatric allergy and immunology : official publication of the European Society of Pediatric Allergy and Immunology. 2010;21(6):895–9.

13. Johansson H, Norlander K, Hedenstrom H, Janson C, Nordang L, Nordvall L, et al. Exercise-induced dyspnea is a problem among the general adolescent population. Respiratory medicine. 2014;108(6):852–8.

14. Christensen PM, Thomsen SF, Rasmussen N, Backer V. Exercise-induced laryngeal obstructions: prevalence and symptoms in the general public. European archives of oto-rhino-laryngology : official journal of the European Federation of Oto-Rhino-Laryngological Societies (EUFOS) : affiliated with the German Society for Oto-Rhino-Laryngology - Head and Neck Surgery. 2011;268(9):1313–9.

15. Shembel AC, Hartnick CJ, Bunting G, Ballif C, Shaiman S, de Guzman V, et al. Perceptual Clinical Features in Exercise-Induced Laryngeal Obstruction (EILO): Toward Improved Diagnostic Approaches. Journal of voice : official journal of the Voice Foundation. 2018.

16. Nielsen EW, Hull JH, Backer V. High prevalence of exercise-induced laryngeal obstruction in athletes. Medicine and science in sports and exercise. 2013;45(11):2030–5.

17. Abu-Hasan M, Tannous B, Weinberger M. Exercise-induced dyspnea in children and adolescents: if not asthma then what? Annals of allergy, asthma & immunology : official publication of the American College of Allergy, Asthma, & Immunology. 2005;94(3):366–71.

18. Khan DA. Exercise-induced bronchoconstriction: burden and prevalence. Allergy and asthma proceedings. 2012;33(1):1–6.

19. Seear M, Wensley D, West N. How accurate is the diagnosis of exercise induced asthma among Vancouver schoolchildren? Archives of disease in childhood. 2005;90(9):898–902.

20. Pedersen ESL, de Jong CCM, Ardura-Garcia C, Barben J, Casaulta C, Frey U, et al. The Swiss Paediatric Airway Cohort (SPAC). ERJ open research. 2018;4(4).

21. Harris PA, Taylor R, Thielke R, Payne J, Gonzalez N, Conde JG. Research electronic data capture (REDCap)--a metadata-driven methodology and workflow process for providing translational research informatics support. Journal of biomedical informatics. 2009;42(2):377–81.

22. von Elm E, Altman DG, Egger M, Pocock SJ, Gotzsche PC, Vandenbroucke JP. The Strengthening the Reporting of Observational Studies in Epidemiology (STROBE) statement: guidelines for reporting observational studies. Journal of clinical epidemiology. 2008;61(4):344–9.

23. Van den Broeck J, Willie D, Younger N. The World Health Organization child growth standards: expected implications for clinical and epidemiological research. European journal of pediatrics. 2009;168(2):247–51.

24. Braun-Fahrlander C, Gassner M, Grize L, Minder CE, Varonier HS, Vuille JC, et al. Comparison of responses to an asthma symptom questionnaire (ISAAC core questions) completed by adolescents and their parents. SCARPOL-Team. Swiss Study on Childhood Allergy and Respiratory Symptoms with respect to Air Pollution. Pediatric pulmonology. 1998;25(3):159–66.

25. Olson LM, Radecki L, Frintner MP, Weiss KB, Korfmacher J, Siegel RM. At what age can children report dependably on their asthma health status? Pediatrics. 2007;119(1):e93–102.

26. Griffin SA, Walsted ES, Hull JH. Breathless athlete: exercise-induced laryngeal obstruction. British journal of sports medicine. 2018;52(18):1211–2.

27. Hall A, Thomas M, Sandhu G, Hull JH. Exercise-induced laryngeal obstruction: a common and overlooked cause of exertional breathlessness. The British journal of general practice : the journal of the Royal College of General Practitioners. 2016;66(650):e683–5.

